# Cannabidiol Modulates Right Fronto-Parietal Connectivity in Autistic Children: A Secondary EEG Analysis from a Randomised Placebo-Controlled Crossover Trial

**DOI:** 10.1101/2025.09.09.25335467

**Authors:** Nina-Francesca Parrella, Aron T. Hill, Peter G. Enticott, Luke A. Downey, Sarah Catchlove, Talitha C. Ford

**Affiliations:** Cognitive Neuroscience Unit, School of Psychology, Deakin University, Burwood, Australia; Centre for Mental Health & Brain Sciences, School of Health Sciences, Swinburne University of Technology, Melbourne, Australia; Eastern Health Clinical School, Monash University, Melbourne, Australia

## Abstract

Autistic children often show altered brain network connectivity, including circuitry implicated in social cognition. Cannabidiol (CBD) has emerged as a potential therapy for autism, but its neural mechanisms remain under-researched. Here, we report an analysis of resting-state electroencephalography (EEG) data collected as part of a double-blind, placebo-controlled crossover trial, to explore whether 12 weeks of oral CBD oil treatment could modulate functional connectivity in autistic children. Twenty-nine autistic children (5-12 years) completed two 12-week intervention periods (CBD and placebo, randomised) separated by an 8-week washout. Before and after each intervention period, five minutes of eyes-open resting-state EEG data were obtained from a subset of 19 participants (8 female). Functional connectivity was calculated across 16 electrode-pairs in the alpha, beta, theta, and gamma bands. Paired t-tests comparing post-versus pre-intervention connectivity values were Benjamini-Hochberg false discovery rate (FDR)-corrected within each band (*p*_FDR_ < .05). Beta-band connectivity between the right frontal and right inferior-parietal electrodes increased following CBD (*p* < .001, *p*_FDR_ = .016) and alpha connectivity showed an increase that did not remain following FDR correction (*p* = .036, *p*_FDR_ = .071). No significant connectivity differences were observed following placebo for any frequency band (*p*_FDR_ > .60). These findings provide preliminary evidence that CBD might enhance connectivity within social cognitive brain networks in autism but will require replication in larger samples.

## Introduction

Autism spectrum disorder (autism) is a neurodevelopmental condition characterised by challenges in social communication, restricted interests, and repetitive behaviours (American Psychiatric Association, 2013). Growing evidence indicates that autism is associated with altered patterns of brain connectivity, including differences in synchronised neural oscillations across cortical regions (Neo et al., 2023). Resting-state EEG studies have found that autistic children can have altered functional connectivity patterns; for example, reduced long-range connectivity between frontal and parietal brain regions relative to typically developing peers (Geng et al., 2023). These connectivity differences are often frequency specific, with some reports describing lower connectivity in the alpha band alongside elevated connectivity in higher-frequency bands (Wantzen et al., 2022).

Converging neuroimaging evidence suggests that these oscillatory alterations reflect atypical maturation of large-scale “social brain” networks that support social attention, communication, and flexible cognitive control in autism (Hong et al., 2019; Schurz et al., 2021). Given that anterior-posterior networks are implicated in attention and social processing (Minnigulova et al., 2025), differences in these regions may underlie social and cognitive difficulties in autism. EEG and magnetoencephalography (MEG) studies have demonstrated that synchrony within right fronto-parietal beta networks supports decoding of social cues and theory-of-mind in typical development (Schurz et al., 2021). Fronto-parietal beta is reduced in autistic cohorts during face- and emotion processing (Safar et al., 2018), and in neurotypical cohorts,autistic trait-variation relates to connectivity within right temporo-parietal networks (Hill et al., 2022). These findings implicate fronto-parietal beta connectivity as a plausible neural marker and potential therapeutic target for social communication in autism.

Cannabidiol (CBD) is a non-intoxicating compound derived from *Cannabis sativa L*. that has gained interest as a potential therapeutic in neurodevelopmental disorders including autism (Cairns et al., 2023; Loss et al., 2021). Preliminary studies suggest CBD may alleviate certain autism-related symptoms (including social functioning and anxiety), although robust evidence is still emerging (Hacohen et al., 2022; Mazza et al., 2024; Parrella et al., 2024). From a mechanistic perspective, research in adults indicates that CBD can alter neural activation and connectivity (Bhattacharyya et al., 2018). In a randomised, placebo-controlled crossover trial, a single 600 mg dose of CBD enhanced resting-state connectivity between the cerebellar vermis and striatum in autistic adults relative to controls (Pretzsch, Voinescu, et al., 2019). Magnetic resonance spectroscopy (MRS) in the same cohort showed that the dose lowered dorsomedial-prefrontal gamma-aminobutyric acid (GABA) more than Glx (glutamate + glutamine), increasing the Glx:GABA ratio for the autistic group, while GABA increased in controls (Pretzsch, Freyberg, et al., 2019). This excitatory shift may explain the strengthened vermis and striatal connectivity and aligns with evidence that higher Glx and lower GABA predicts stronger resting-state connectivity (Stagg et al., 2014).

EEG connectivity studies of CBD are limited and mainly from refractory epilepsy cohorts (Anderson et al., 2020; Armstrong et al., 2022; Grayson et al., 2021; Morales Chacón et al., 2021). Using coherence, synchronisation-likelihood and graph measures, these studies suggest CBD can shift large-scale networks, with increased beta and sometimes alpha efficiency/modularity (Anderson et al., 2020) and band-specific changes in synchrony (Armstrong et al., 2022; Grayson et al., 2021; Morales Chacón et al., 2021). EEG-connectivity studies examining the effects of CBD in autism have not yet been conducted, highlighting the rationale for the present investigation. The proposed mechanisms of CBD involve modulation of endocannabinoid, GABAergic, and glutamatergic systems which are likely to influence oscillatory brain dynamics (Aychman et al., 2023). Regional levels of GABA and glutamate are strong determinants of network-level connectivity, suggesting that the effects of CBD on this neurochemical balance may translate into altered connectivity in autism (Kapogiannis et al., 2013; Kwon et al., 2014). Considering these studies, it is possible that CBD could modulate atypical neural connectivity in autism, and that this might be one mechanism that underpins clinical benefits.

The present study reports analysis of the EEG data collected as part of our randomised, double-blind, placebo-controlled crossover trial of oral CBD oil in autistic children (Parrella et al., 2024). While we have previously reported clinical outcomes for the trial, this paper is the first report of the associated EEG data. Here we examined whether the 12-week CBD oil intervention modulated functional connectivity across a broad range of EEG frequencies (delta [1-4 Hz], theta [4-8 Hz], alpha [8-13 Hz], beta [13-30 Hz], and gamma [30-45 Hz]). We analysed a predefined set of 16 electrode pairs, including anterior-posterior and fronto-parietal pathways relevant to social cognition (full list provided in the Methods section). The selected electrode pairs are relevant to neurodevelopmental research and have previously been implicated in traits associated with autism (Aykan et al., 2021; Hill et al., 2022). We also explored whether connectivity changes relate to concurrent changes in social responsiveness and behavioural/emotional functioning. We hypothesised that the CBD intervention would increase low-frequency functional connectivity between frontal and temporo-parietal areas (i.e., social brain regions including medial prefrontal cortex (mPFC), anterior cingulate cortex (ACC), and temporoparietal junction/posterior superior temporal sulcus (TPJ/pSTS) (Frith & Frith, 2007), and that these increases would be associated with improvements in social communication outcomes.

## Materials and Methods

### Study Design

This paper presents analysis of EEG data collected from a randomised, double-blind, placebo-controlled crossover trial (ACTRN12622000437763; behavioural outcomes reported previously in Parrella et al., 2024), which was approved by the Deakin University Human Research Ethics Committee (DUHREC 2020-071) and conducted in accordance with the Declaration of Helsinki. Written informed consent was obtained from parents/guardians, and children provided assent. Participants underwent two 12-week intervention periods (CBD oil and placebo) separated by an 8-week washout. CBD was administered orally as Medigrowth CBD100 oil (containing cannabis-derived terpenes), starting at 5 mg/kg/day and titrated up to 10 mg/kg/day. The placebo oil was matched in appearance and taste. Parents or guardians administered the oil at home, and adherence was monitored via a daily treatment log. Treatment log details and adverse events are reported in Parrella et al. (2024).

### Participants

Thirty-four children (aged 5-12 years, 11 female) with an existing diagnosis of autism spectrum disorder (ASD) were enrolled in the study (Table 1). ASD diagnoses were confirmed by qualified clinicians using standard diagnostic instruments (e.g., Autism Diagnostic Observation Schedule, parent interview) and review of medical reports. All participants had no changes to psychotropic medication during the study and did not have any identified genetic or chromosomal disorders. Of the 34 children enrolled, 29 completed the trial, and 19 underwent EEG recording (baseline and post-intervention visits, age 5-12 years, 8 female; see Table 1). EEG could not be obtained from all enrolled children because some chose not to undergo EEG procedures, and others could not tolerate EEG procedures due to behavioural disruption and/or sensory sensitivities (see CONSORT diagram in Figure 1).

**Figure 1.**
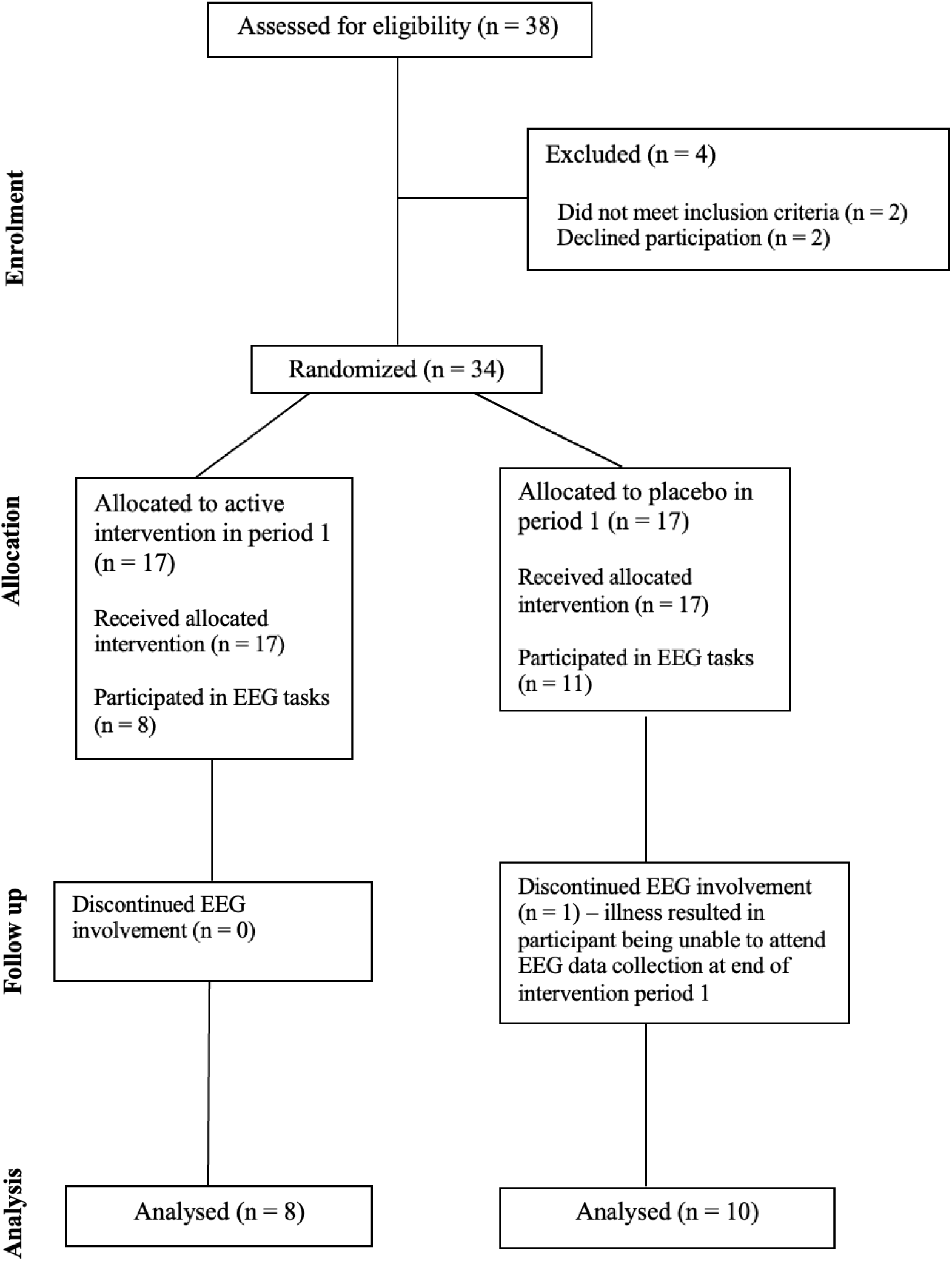
CONSORT Diagram depicting participants completing EEG component of trial.

**Table 1.**
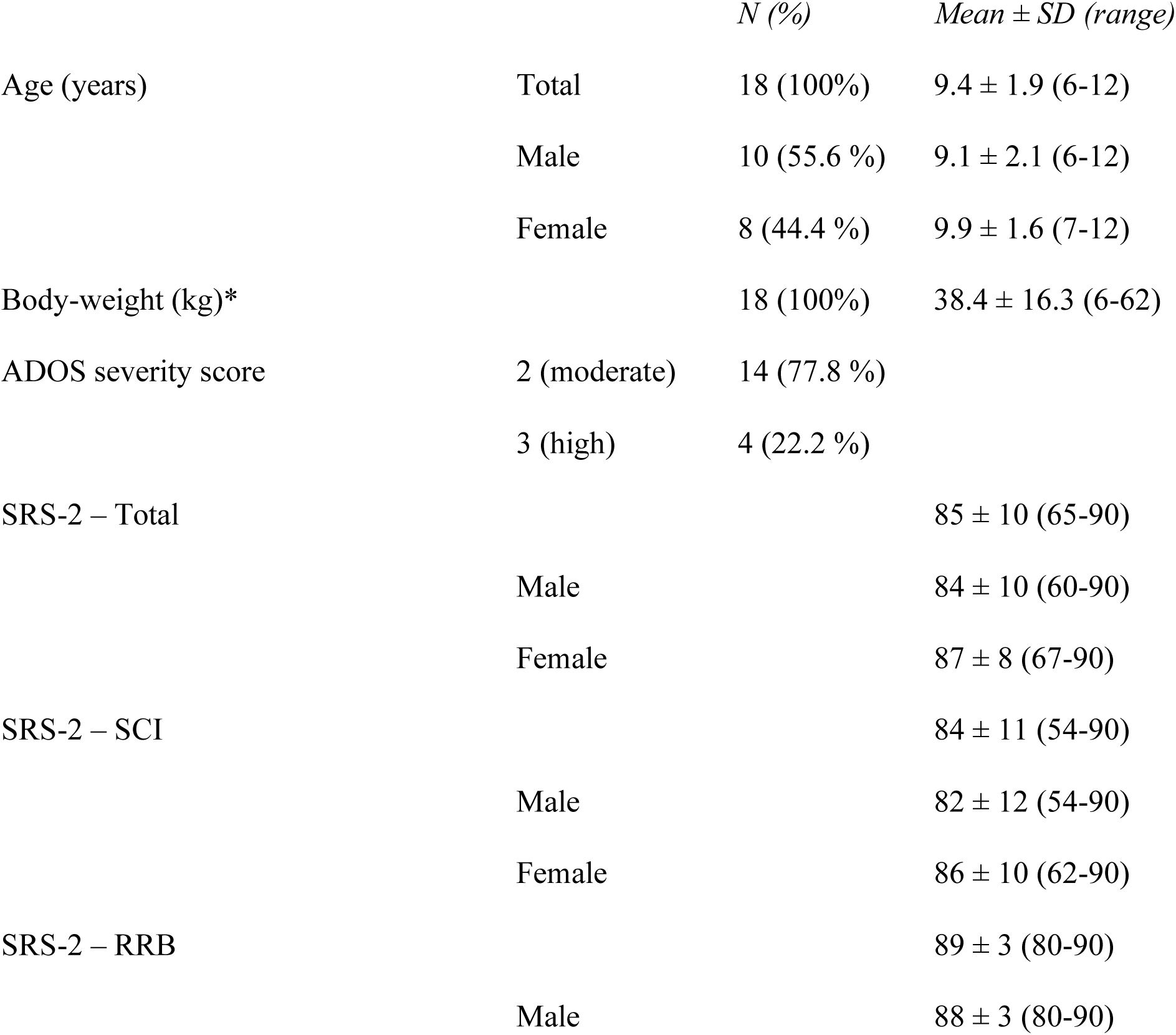
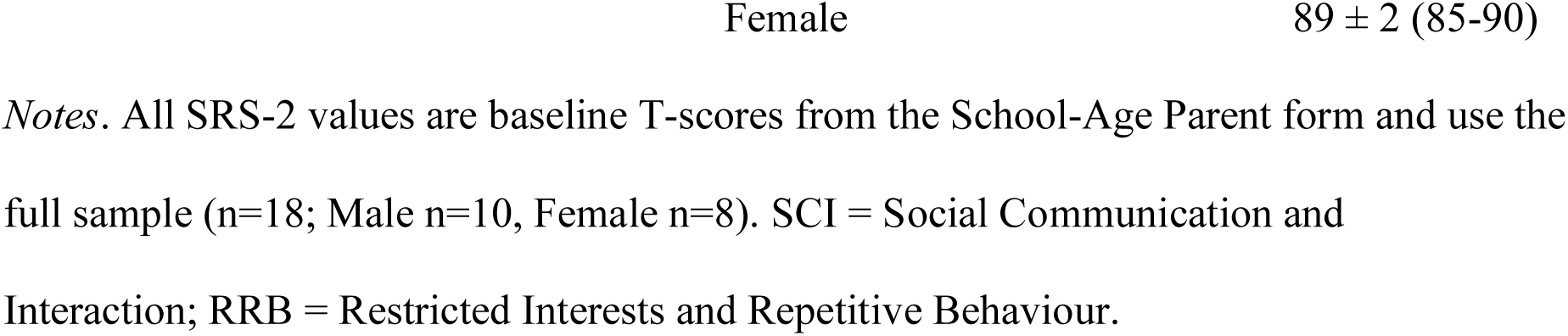
Demographic characteristics and baseline Social Responsiveness Scale-2 (SRS-2) T-scores.

### EEG Acquisition and Preprocessing

Resting-state EEG was recorded before and after each 12-week intervention period. Post-treatment recording sessions took place within five days of the planned 12-week (84-day) endpoint; however, to ensure EEG always occurred within 24 hours of the last CBD/placebo dose, participants continued taking the assigned dosage until the day before (or morning of) their EEG visit. Due to participant availability, the dosing period therefore ranged from 81 to 89 days (M = 85, SD = 2.3).

Data were acquired using a 64-channel HydroCel Geodesic Sensor Net (Electrical Geodesics, Inc., Eugene OR, USA). Each electrode consisted of an Ag/AgCl sensor surrounded by an electrolyte-wetted sponge, and impedances were kept below ∼50 kΩ for the duration of recording. Signals were amplified using a NetAmps 400 amplifier and digitised at 1000 Hz with an online reference at the vertex (Cz). Participants sat quietly in a dimly lit room with eyes open for 5 minutes while EEG was recorded. They were instructed to fixate on a cross and minimise movement. Clinically trained research staff monitored the children to ensure they remained calm, content, and relatively still.

EEG data were pre-processed in MATLAB (R2021a; The Mathworks, Massachusetts, USA) using the EEGLAB toolbox (v2023.0) and custom scripts. The Reduction of Electroencephalographic Artifacts (RELAX-Jr) software was employed to clean each EEG dataset. RELAX-Jr software is a pipeline specifically developed for pre-processing of EEG data collected from neurodevelopmental populations (Hill et al., 2024). RELAX-Jr utilises empirical methods to identify and mitigate non-neural artifacts, incorporating multi-channel Wiener filters and wavelet-enhanced independent component analysis (ICA). Data were bandpass filtered between 0.25 and 80 Hz using a fourth-order zero-phase Butterworth filter and a notch filter (47-53 Hz) was applied to eliminate line noise. Noisy channels were identified and removed through a multi-step process using the ‘findNoisyChannels’ function from the PREP pipeline (Bigdely-Shamlo et al., 2015). Multi-channel Wiener filtering (Somers et al., 2018) was then applied to clean blinks, muscle activity, and horizontal eye movement, followed by robust average re-referencing and wavelet-enhanced ICA. Components for cleaning were identified using the adjusted-ADJUST automated independent component classification algorithm designed for Geodesic sensor nets (Leach et al., 2020). Rejected electrodes were interpolated back into the dataset using spherical interpolation, and all pre-processed data files were visually inspected prior to further analysis.

### Connectivity Analysis

Functional connectivity was quantified across 16 predefined (*a priori*) electrode pairs selected from previous EEG connectivity work (Aykan et al., 2021; Hill et al., 2022) to capture long-distance “social-brain” pathways: right anterior (F4–T8, C4–T8), left anterior (F3–T7, C3–T7), right posterior (C4–P8, P4–T8), left posterior (C3–P7, P3–T7), right fronto-parietal (F4–P4, F4–P8, F8–P4, F8–O8), and left fronto-parietal (F3–P3, F3–P7, F7–P3, F7– P7). For each participant and session, the cleaned EEG data were first re-referenced using a surface Laplacian (current-source density) spatial filter to minimise volume-conduction effects (Srinivasan et al., 2007). Weighted phase-lag index (wPLI) connectivity was then computed for the delta (1–4 Hz), theta (4–8 Hz), alpha (8–13 Hz), beta (13–30 Hz), and gamma (30–45 Hz) bands. Connectivity between each electrode pair was extracted for each frequency band using the wPLI with the fieldtrip MATLAB toolbox (Oostenveld et al., 2011). The wPLI connectivity estimate is robust to volume conduction as it disregards instantaneous (zero phase-lag) connections (Vinck et al., 2011). For each frequency band and electrode pair, post-intervention connectivity was compared with its own baseline value using a paired-samples design (CBD and placebo blocks analysed separately). Within-subject difference in connectivity (Post-Pre) was tested for each electrode-band combination. An identical procedure was applied to the placebo condition data for comparison.

### Behavioural measures

This study is part of a larger investigation into changes in clinical outcomes following CBD, and as such, we sought to examine whether the changes in clinical outcomes reported in Parrella et al. (2024) were associated with any change in functional connectivity identified in this study. Parent-reported behavioural measures included here are: the Social Responsiveness Scale-2 (SRS-2; Constantino & Gruber, 2012), Developmental Behaviour Checklist-2 (DBC-2; Einfeld & Tonge, 2002), the Behavior Rating Inventory of Executive Function (BRIEF; Gioia et al., 2015), and the Vineland Adaptive Behavior Scales, Third Edition (Vineland-3; Sparrow et al., 2016).

We calculated change scores for these behavioural measures during the CBD phase (e.g. baseline SRS-2 – post-CBD SRS-2, so that positive change reflects improvement/reduction in symptom severity on the SRS-2). These behavioural change scores were used in exploratory correlational analyses to investigate associations with EEG connectivity changes.

### Statistical Analysis

#### Connectivity

For each electrode pair and frequency band, a paired-samples *t*-test compared the post-intervention connectivity value with the corresponding baseline value for that same treatment condition (CBD and placebo analysed separately). This yielded one *t*-statistic per electrode pair per frequency band. Normality of the paired differences (Post-Pre) was assessed for all 80 electrode x band combinations with the Shapiro-Wilk test (smallest W = 0.92, *p* = .21) and confirmed by visual inspection of Q-Q plots; all distributions met the normality assumption, thus parametric paired-samples *t*-tests were used; significance was defined at *p* < .05 (two-tailed). To control for multiple comparisons, false discovery rate (FDR) correction was applied separately within each frequency band (Benjamini-Hochberg procedure; *p_FDR_* < 0.05). The placebo data were analysed using the same approach. Effect sizes for significant connectivity differences were calculated as Cohen’s *d* for paired samples. All statistical analyses were performed using MATLAB and Jamovi.

#### Bayesian analysis (primary connection)

To further assess the non-trivial placebo change, we conducted a Bayesian repeated-measures ANOVA for the *a priori* primary connection (F4-P8, beta) with within-subject factors Time (Pre, Post) and Condition (CBD, Placebo), targeting the Time × Condition interaction. We report the posterior estimate of the interaction, its 95% credible interval, and Pr (effect>0). Bayesian analyses were performed in Jamovi (jsq, Bayesian RM ANOVA).

#### Brain-behaviour correlations (exploratory)

For each electrode-frequency band pair showing a significant pre-post CBD effect, we investigated associations with pre-post CBD change in behavioural outcomes (SRS-2, DBC-2, BRIEF, Vineland-3) using Pearson’s r correlation coefficients. Given the exploratory nature and modest sample size, we report *p*-values without correction alongside corrected *p*_FDR_-corrected values and interpret these correlation results cautiously.

## Results

### Participant characteristics

EEG data were collected from 18 participants (see Table 1). All children had moderate-high support needs, 27% showed significant speech delays/differences, and the remainder used fluent speech. Baseline SRS-2 scores averaged in the severe range (T-score > 70), indicating significant social impairment.

### Connectivity change for CBD condition

CBD treatment produced significant pre- to post-intervention increases in F4-P8 connectivity for the beta band (*t*(17) = -3.81, *p* = .001), with a strong effect size (Cohen’s *d* = 0.90), and a smaller increase in the alpha band (*t*(17) = -2.28, *p* = .036, *d* = 0.54). After FDR correction for multiple comparisons within each frequency band, only the beta-band effect remained significant (beta: *p*_FDR_ = .003; alpha: *p*_FDR_ = .071, see Table 2 and Figure 2). No other electrode pair comparisons reached significance after FDR correction in the CBD condition (*p_FDR_* > .05).

**Figure 2.**
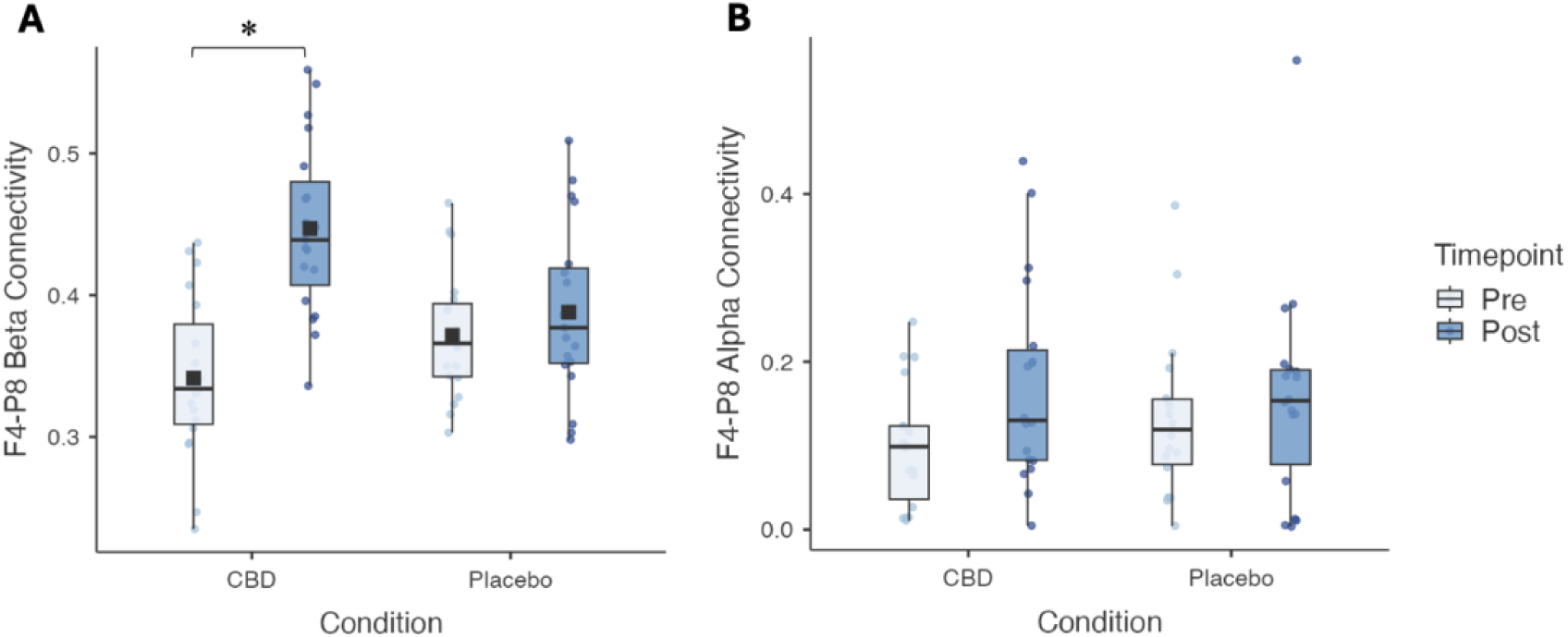
**(A)** Beta band (13–30 Hz): pre- vs post-intervention in cannabidiol (CBD) and placebo periods. **(B)** Alpha band (8–13 Hz): pre- vs post-intervention in CBD and placebo periods. Boxes show median and interquartile range; individual participants are overlaid as points. * denotes significant pre-post change (*p*_FDR_ = .003).

**Figure 3.**
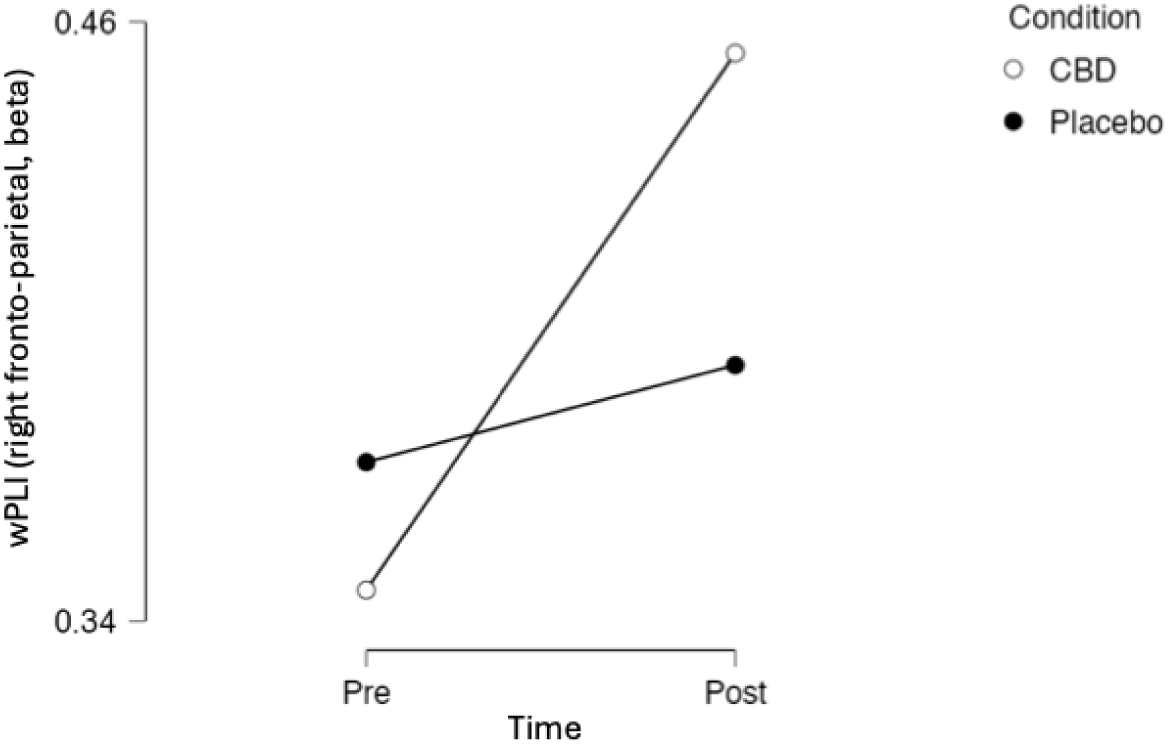
Bayesian estimated marginal means (95% credible intervals) for right fronto-parietal (F4-P8) beta connectivity. Time x Condition supported (BFinclusion = 19.24); cannabidiol (CBD)-placebo difference-in-differences = 0.089 wPLI (95% CrI [L, U].

**Table 2.** Paired t-test results for F4-P8 connectivity (pre-post) in CBD and placebo conditions.

| Condition | Band | *t*-statistic | *df* | *p*-value | *p*<sub>FDR</sub>-value | Cohen's *d* |
| --- | --- | --- | --- | --- | --- | --- |
| CBD | Delta | -1.98 | 17 | 0.065 | 0.129 | 0.47 |
|  | Theta | -0.41 | 17 | 0.688 | 0.688 | 0.10 |
|  | Alpha | -2.28 | 17 | **0.036** | 0.071 | 0.54 |
|  | Beta | -3.81 | 17 | **0.001** | **0.003** | 0.90 |
|  | Gamma | -2.03 | 17 | 0.058 | 0.117 | 0.48 |
| Placebo | Delta | 0.12 | 17 | 0.904 | 0.904 | 0.03 |
|  | Theta | 0.41 | 17 | 0.683 | 0.688 | 0.10 |
|  | Alpha | -0.72 | 17 | 0.482 | 0.482 | 0.17 |
|  | Beta | 0.18 | 17 | 0.859 | 0.859 | 0.04 |
|  | Gamma | 0.99 | 17 | 0.338 | 0.338 | 0.23 |

Notes: CBD = cannabidiol; FDR = false discovery rate; *p*<sub>FDR</sub> = FDR-adjusted *p*-value; *df* = degrees of freedom.

### Connectivity change for placebo condition

In the placebo condition, two electrode pairs showed significant increases prior to correction (F4-T8 beta band, t(17)=2.89, *p* = .010, Cohen’s *d* = 0.68; and F3-P7 in the alpha band, t(17)=-2.18, *p* = .044, Cohen’s *d* = -0.51), but these did not survive FDR adjustment (*p*_FDR_ = .163 and *p*_FDR_ = .351, respectively). Therefore, no reliable differences in connectivity were observed under the placebo condition (see Table 2 and Figure 2).

### Bayesian analysis for F4-P8, beta

A Bayesian repeated-measures ANOVA supported a CBD-specific pre- to post-intervention increase in connectivity. The model including Time, Condition and their interaction was favoured over the null (BF_10_ = 1735), with strong evidence for the Time x Condition effect (BF_inclusion_ = 19.24). The pre- to post-intervention change was +0.108 under CBD and +0.019 under placebo. The difference-in-differences (Bayesian posterior) was 0.089 wPLI (95% credible interval [L, U]; see Table 3).

**Table 3.** Bayesian repeated-measures ANOVA at the right fronto-parietal beta connection.

| Effect | Prior $P(incl)$ | Posterior $P(incl\vert data)$ | $BF_{Inclusion}$ |
| --- | --- | --- | --- |
| Time | 0.60 | 0.999 | 980.89 |
| Condition | 0.60 | 0.884 | 5.08 |
| Time x Condition | 0.20 | 0.828 | 19.24 |

*Notes.* Effects are Time (Pre vs Post), Condition (Cannabidiol; CBD vs Placebo), and Time x Condition (CBD-specific change). $P(incl)$ is the prior inclusion probability and $P(incl|data)$ is the posterior inclusion probability. $BF_{inclusion} > 1$ favours including the effect. Outcome is F4-P8 connectivity quantified by the weighted phase-lag index (wPLI), with participants as the subject factor.

### Connectivity and behavioural measures

Exploratory correlations between connectivity and behavioural outcomes (*n*=17, CBD condition) showed no effects surviving FDR correction (Supplementary Table S1). We focused on F4-P8 connectivity in the alpha band (motivated by prior autism literature on alpha-band connectivity (Safar et al., 2018), and trend level-effect observed for alpha) and in the beta band (given a prominent beta-band connectivity effect observed in our data). All correlations are summarised in Supplementary Table S1; several uncorrected associations were observed (*p*s < .05), but none remained following FDR correction (all *p*_FDR_ > .05). For example, change in F4-P8 alpha connectivity correlated with the SRS-2 Social Communication Index (*r* = 0.51, *p* = 0.046, *p*_FDR_ = 0.31), and change in F4-P8 beta connectivity correlated with Vineland Socialisation (*r* = 0.51, *p* = 0.044, *p*_FDR_ = 0.71). Correlations for the placebo condition were not examined, as there were no significant changes observed in either behavioural outcomes or EEG connectivity measures during the placebo phase.

## Discussion

This randomised crossover trial is among the first to use EEG to investigate the impact of 12 weeks of daily CBD oil on cortical functional connectivity in autistic children. CBD significantly increased beta-band connectivity between right frontal (F4) and right inferior-parietal (P8) electrodes relative to baseline, but not other electrode pairs after FDR correction, and there were no significant changes in connectivity under placebo. This increase in right fronto-parietal beta connectivity was somewhat associated with behavioural change scores (SRS-2 and Vineland-3), although none withstood correction. Given that beta synchrony supports long-range, top-down control of attention in humans (Morillon & Baillet, 2017; Spitzer & Haegens, 2017), the observed fronto-parietal increase may reflect strengthened communication within networks relevant to social cognition.

CBD has been shown to modulate GABA and glutamate neurotransmission, which alters cortical excitation/inhibition (E/I) balance (Pretzsch, Freyberg, et al., 2019). Altered E/I balance has also been implicated in autism, and social processing difficulties in particular (Canitano & Pallagrosi, 2017; Cochran et al., 2015; Ford & Crewther, 2014; Gaetz et al., 2014). While GABA modulation shows promise for addressing core social difficulties in autism (Veenstra-VanderWeele et al., 2017), there are currently no approved pharmacological therapeutics that target this domain, with strong reliance on behavioural interventions. The influence of CBD on E/I mechanisms therefore supports the hypothesis that social processing might be effectively modulated by CBD intervention (Pretzsch, Freyberg, et al., 2019). Consistent with our CBD-related increase in right fronto-parietal beta, EEG connectivity studies of CBD, though limited and mainly from refractory epilepsy cohorts, have suggested that CBD can shift large-scale networks. Increased beta and sometimes alpha efficiency/modularity have been reported (Anderson et al., 2020), as well as band-specific changes in synchrony (Armstrong et al., 2022; Grayson et al., 2021; Morales Chacón et al., 2021).

Beta-band oscillatory activity, which supports long-range, top-down communication, is sensitive to GABAergic modulation and has been used as a marker of cortical GABA function (Groth et al., 2021; Spitzer & Haegens, 2017; Teixeira et al., 2021). This sensitivity may underlie the CBD-related increase in right fronto-parietal beta connectivity. High-beta (20-30 Hz) connectivity is considered an EEG marker of GABAergic inhibitory tone (Barone & Rossiter, 2021; Baumgarten et al., 2016; Groth et al., 2021). In chronic pain cohorts, reduced beta connectivity, particularly in the left basolateral amygdala, has been associated with more severe affective pain symptoms (Makowka et al., 2023). Independent connectomic work shows that the strength of cortical beta networks covaries positively with regional dopamine-receptor and transporter density, which suggests dopaminergic contribution to large-scale beta oscillations (Chikermane et al., 2024).

The increase in beta connectivity observed in this study may indicate a modest normalisation of inhibitory tone for these autistic children. This is notable given that right fronto-parietal networks are implicated in social cognition and are often under-connected in autism (Lombardo et al., 2011; Safar et al., 2018; Wantzen et al., 2022). Hypo-connectivity of fronto-parietal beta rhythms has been reported in autistic children during rest and social-cognitive tasks (e.g., during face processing) and is thought to contribute to atypical attention and social information integration (Safar et al., 2018; Wantzen et al., 2022). MEG/EEG studies show reduced fronto-posterior beta synchrony during social-cognitive tasks (e.g., emotional face processing; name recognition) in autistic adolescents/young adults (Nowicka et al., 2016; Safar et al., 2021).

Neuroimaging research also implicates right-lateralised temporo-parietal (rTPJ/pSTS) and fronto-parietal networks in social cognition, with atypical engagement/connectivity reported in autism (Lombardo et al., 2011; Saxe & Kanwisher, 2003). Large multi-cohort and meta-analytic studies associate rTPJ-centred and fronto-parietal connectivity abnormalities with social symptom severity in autism, and separate the rTPJ into social and attentional subregions that are implicated in social-communication (Hao et al., 2022; Holiga et al., 2019). Meta-analytic EEG/MEG work indicates that individuals with autism show long-range underconnectivity most evident in lower-frequency bands and atypical hemispheric lateralisation (O’Reilly et al., 2017). Variation in right-lateralised temporo-/fronto-parietal connectivity may covary with social communication differences in autism. Evidence for another frequency band implicates right-lateralised networks in neurotypical samples, with theta connectivity predicting higher autistic-trait scores (Aykan et al., 2021; Hill et al., 2022), suggesting these networks are behaviourally relevant to autism and social communication.

One interpretation of the current findings is that children with weaker fronto-parietal beta synchrony may have shifted toward a level more characteristic of typical functional connectivity following treatment with CBD. This is supported by pharmacological evidence that CBD can elevate cortical GABA levels (Pretzsch, Freyberg, et al., 2019; Ruffolo et al., 2022). However, this interpretation remains speculative without a neurotypical comparison group. The right fronto-parietal (F4-P8) beta-band post-treatment connectivity increase in the present autistic cohort suggests that CBD may strengthen long-range communication and potentially support under-connected network activity. We also observed a modest rise in alpha connectivity at the same electrode pair. Although this did not survive correction, the direction aligns with our beta-band findings as well as reports in neurotypical children that have shown reduced anterior-posterior alpha synchrony, which supports integrated attention and social perception (Hill et al., 2025). Complementary EEG-microstate research has shown atypical alpha-microstate dynamics in autistic adults, suggesting disruption to large-scale alpha networks (Das et al., 2024). These observations indicate that larger samples will be needed to determine whether CBD can reliably modulate alpha-band communication, as well as beta.

Social functioning in autism relies on coordination between frontal regions that support social decision-making and theory-of-mind, and posterior perceptual regions that decode faces and social language (Pitcher & Ungerleider, 2021; Vukovic & Shtyrov, 2017). Extant EEG and MEG research shows that the integrity of right-lateralised fronto-parietal beta networks predicts decoding of social cues and theory-of-mind accuracy in typically developing children (Schurz et al., 2021) and is attenuated in autistic samples during face or emotion viewing (Safar et al., 2018). CBD-related increases in right fronto-parietal beta synchrony likely reflects more efficient communication between posterior-frontal communication and may support improved social functioning in autism.

In line with this premise, exploratory analyses revealed trend-level associations: greater alpha-band increases were linked to improved SRS-2 Social Cognition scores, and larger beta-band gains were associated with higher Vineland Socialisation scores. These tentative findings would align with reports that fronto-parietal alpha/beta-band connectivity indicates social-communication in autism (Das et al., 2024; Mathewson et al., 2012; Safar et al., 2022). However, likely due to limited statistical power, none survived multiple-comparison correction. Larger samples are needed to clarify whether connectivity change mediates CBD-related behavioural benefits.

### Limitations and future directions

There are several limitations to consider when interpreting these findings. First, the number of participants with usable EEG data was small (*n*=18), limiting statistical power and generalisability of results. While the F4-P8 beta-band increase was robust and survived correction for multiple comparisons, it is possible that the small sample reduced detection of additional CBD-related changes. Although the crossover design reduces between-participant variance, residual order or carryover effects cannot be ruled out and should be monitored in larger samples. Second, we analysed resting-state EEG (eyes open). While this provides important information on spontaneous network activity, task-based paradigms that engage social-cognitive processing might provide additional insight into specific networks of interest in autism, although acquiring artefact-free task data in this cohort is challenging.

Third, methodologically, we focused on *a priori* electrode pairs and standard oscillatory frequency bands. This limits the number of tests but may potentially miss more distributed network effects. Fourth, the study did not include a neurotypical comparison group. Without a normative reference group, we cannot determine whether baseline connectivity (or its modulation by CBD) differs from age-matched neurotypical profiles, nor whether any normalisation toward typical connectivity occurs. Including a matched neurotypical group would clarify autism-specificity and contextualise the magnitude of change.

Fifth, it would be informative to investigate if the changes in EEG connectivity are maintained beyond the immediate intervention period. Longitudinal follow-up could test maintenance or waning of effects. Sixth, this study evaluated chronic CBD/placebo exposure with titration to a stable daily dose. Single-dose (acute) studies and dose-response designs could clarify whether lower or higher doses produce different EEG effects and their time course. Finally, exploring EEG connectivity in relation to pharmacokinetic data (e.g. CBD plasma levels) might also help understand individual variability. It would be worth exploring the extent to which baseline EEG features predict which children benefit most from CBD. For example, whether children with lower baseline beta connectivity show larger increases and greater social improvement.

### Conclusion

This analysis of EEG data from a placebo-controlled crossover trial of oral CBD in autistic children demonstrated that daily administration of CBD oil for 12 weeks enhanced right fronto-parietal functional connectivity in the beta-band. This change was accompanied by improvements in some behavioural domains (social relating, anxiety, and parental stress, reported in Parrella et al., (2024), suggesting a potential link between the effect of CBD on the brain and clinical outcomes, however such links were not supported in this study. While preliminary, these findings offer initial evidence that CBD may improve brain connectivity patterns in autistic children. This work demonstrates the feasibility and value of incorporating EEG measures into autism clinical trials to investigate the underlying neurophysiological mechanisms associated with behavioural outcomes.

## Data Availability

De-identified EEG and behavioural data, along with analysis scripts, are available on reasonable request, subject to ethics approval and data-sharing agreements.

