## Supplemental Material 1 for "Cannabidiol Modulates Right Fronto-Parietal Connectivity in Autistic Children: A Secondary EEG Analysis from a Randomised Placebo-Controlled Crossover Trial"

***Table S1.*** Correlations between alpha and beta F4-P8 electron pair and behavioural outcomes.

| | Alpha | | | Beta | | |
|---|---|---|---|---|---|---|
| Behavioural measure | *r* | *p* | $p_{FDR}$ | *r* | *p* | $p_{FDR}$ |
| SRS-2: Awareness | 0.26 | 0.329 | 0.57 | -0.14 | 0.608 | 0.95 |
| SRS-2: Cognition | **0.56** | **0.023** | **0.21** | 0.04 | 0.876 | 0.97 |
| SRS-2: Communication | 0.46 | 0.070 | 0.31 | -0.41 | 0.113 | 0.78 |
| SRS-2: Motivation | 0.42 | 0.107 | 0.32 | -0.02 | 0.942 | 0.97 |
| SRS-2: Restricted/Repetitive | 0.03 | 0.900 | 0.94 | -0.16 | 0.544 | 0.95 |
| SRS-2: Social Communication Index | **0.51** | **0.046** | **0.31** | -0.19 | 0.470 | 0.95 |
| SRS-2: Total | 0.43 | 0.097 | 0.32 | -0.20 | 0.459 | 0.95 |
| BRIEF: Organization of Materials | **0.65** | **0.006** | **0.17** | 0.17 | 0.530 | 0.95 |
| BRIEF: Task-Monitor | 0.33 | 0.207 | 0.40 | 0.12 | 0.666 | 0.95 |
| BRIEF: Plan/Organize | 0.34 | 0.194 | 0.40 | 0.05 | 0.860 | 0.97 |
| BRIEF: Working Memory | 0.17 | 0.527 | 0.75 | 0.33 | 0.211 | 0.78 |

| | | | | | | |
|---|---|---|---|---|---|---|
| BRIEF: Initiate | 0.26 | 0.337 | 0.57 | 0.35 | 0.179 | 0.78 |
| BRIEF: Emotional Control | 0.34 | 0.193 | 0.40 | -0.01 | 0.971 | 0.97 |
| BRIEF: Shift | 0.38 | 0.149 | 0.40 | -0.04 | 0.894 | 0.97 |
| BRIEF: Self-Monitor | 0.05 | 0.859 | 0.94 | -0.05 | 0.850 | 0.97 |
| BRIEF: Inhibit | 0.13 | 0.635 | 0.84 | 0.08 | 0.758 | 0.97 |
| DBC-2: Total | 0.35 | 0.185 | 0.40 | -0.12 | 0.654 | 0.95 |
| DBC-2: Disruptive | **0.59** | **0.016** | **0.21** | 0.12 | 0.649 | 0.95 |
| DBC-2: Self-Absorbed | 0.12 | 0.654 | 0.84 | -0.18 | 0.496 | 0.95 |
| DBC-2: Communication Disturbance | 0.22 | 0.409 | 0.65 | -0.32 | 0.231 | 0.78 |
| DBC-2: Anxiety | 0.47 | 0.067 | 0.31 | 0.03 | 0.922 | 0.97 |
| DBC-2: Social-Relating | 0.03 | 0.923 | 0.94 | -0.22 | 0.413 | 0.95 |
| Vineland-3: Adaptive Behavior Composite | 0.07 | 0.801 | 0.94 | 0.49 | 0.053 | 0.71 |
| Vineland-3: Communication | 0.19 | 0.484 | 0.73 | 0.36 | 0.176 | 0.78 |
| Vineland-3: Daily Living Skills | 0.10 | 0.716 | 0.88 | 0.33 | 0.205 | 0.78 |

| | | | | | | |
|---|---|---|---|---|---|---|
| Vineland-3: Socialization | 0.02 | 0.943 | 0.94 | **0.51** | **0.044** | **0.71** |
| Vineland-3: Motor Skills | 0.81 | 0.098 | 0.32 | 0.61 | 0.274 | 0.82 |

*Notes:* SRS-2 = Social Responsiveness Scale, Second Edition; BRIEF = Behavior Rating Inventory of Executive Function; DBC-2 = Developmental Behaviour Checklist, version 2; Vineland-3 = Vineland Adaptive Behavior Scales, Third Edition; r = Pearson correlation coefficient; $p$ = uncorrected p-value; $p_{FDR}$ = FDR-adjusted p-value.
